# A systematic review of early neuroimaging and neurophysiological biomarkers for post-stroke mobility prognostication

**DOI:** 10.1101/2025.09.22.25336034

**Authors:** Cristina Levy, Emily J Dalton, Jennifer K. Ferris, Bruce C.V. Campbell, Amy Brodtmann, Sandra Brauer, Leonid Churilov, Kathryn S Hayward

**Author notes:** E.J. Dalton and J.K. Ferris contributed equally. **Corresponding Author** Kate Hayward, PhD, University of Melbourne, Department of Physiotherapy, Level 7 Alan Gilbert Building, 161 Barry Street Parkville, Victoria 3010, Australia.

## Abstract

**Background:** Accurate prognostication of mobility outcomes is essential to guide rehabilitation and manage patient expectations. The prognostic utility of neuroimaging and neurophysiological biomarkers remains uncertain when measured early post-stroke. This systematic review aimed to examine the prognostic capacity of early neuroimaging and neurophysiological biomarkers of mobility outcomes up to 24-months post-stroke.

**Methods:** MEDLINE and EMBASE were searched from inception to June 2025. Cohort studies that reported neuroimaging or neurophysiological biomarkers measured ≤14-days post-stroke and mobility outcome(s) assessed >14-days and ≤24-months post-stroke were included. Biomarker analyses were classified by statistical analysis approach (association, discrimination/classification or validation). Magnitude of relevant statistical measures was used as the primary indicator of prognostic capacity. Risk of bias was assessed using the Quality in Prognostic Studies tool. Meta-analysis was not performed due to heterogeneity.

**Results:** Twenty reports from 18 independent study samples (n=2,160 participants) were included. Biomarkers were measured a median 7.5-days post-stroke, and outcomes were assessed between 1- and 12-months. Eighty-six biomarker analyses were identified (61 neuroimaging, 25 neurophysiological) and the majority used an association approach (88%). Few used discrimination/classification methods (11%), and only one conducted internal validation (1%); an MRI-based machine learning model which demonstrated excellent discrimination but still requires external validation. Structural and functional corticospinal tract integrity were frequently investigated, and most associations were small or non-significant. Lesion location and size were also commonly examined, but findings were inconsistent and often lacked magnitude reporting. Methodological limitations were common, including small sample sizes, moderate to high risk of bias, poor reporting of magnitudes, and heterogeneous outcome measures and follow-up time points.

**Conclusions:** Current evidence provides limited support for early neuroimaging and neurophysiological biomarkers to prognosticate post-stroke mobility outcomes. Most analyses remain at the association stage, with minimal progress toward validation and clinical implementation. Advancing the field requires international collaboration using harmonized methodologies, standardised statistical reporting, and consistent outcome measures and timepoints.

**Registration:** URL: https://www.crd.york.ac.uk/prospero/; Unique identifier: CRD42022350771.

## Introduction

Two-thirds of individuals have difficulty walking independently early post-stroke and this limitation persists at three months for many^1,2^. The ability to mobilize after stroke is consistently identified by stroke survivors as a key rehabilitation goal and top research priority^3^. Early insights into who will regain independent mobility, including transfers and walking, is critical to guide rehabilitation planning, goal setting and communication of expectations with patients and families. Inaccurate prognostication can lead to suboptimal patient outcomes, misaligned expectations, and inefficient use of healthcare resources.

Prognostication of mobility outcomes remains a significant challenge, despite its clinical importance. Prognostic estimates are often inaccurate, even among highly experienced clinicians^4^. In clinical practice, prognostication is typically informed by clinical judgement based on early bedside observation and routine clinical measures^4^. However, stroke recovery is heterogeneous, and individuals with similar initial impairments can experience markedly different long-term outcomes^1,2^. This variability limits the prognostic utility of clinical measures alone and suggests a need for indicators that better account for the multifactorial and biologically complex nature of post-stroke recovery.

Stroke recovery biomarkers may improve the accuracy of prognosticating mobility. A stroke recovery biomarker is an indicator of disease state that measures underlying molecular and cellular processes that may be difficult to measure directly and can be used to understand outcome or predict recovery or treatment response^5^. In 2017, the Stroke Recovery and Rehabilitation Roundtable (SRRR)^5^ identified biomarkers as a key research priority. While substantial progress has been made to identify upper limb recovery biomarkers, comparatively little research has focused on mobility.

Previous systematic reviews^6–8^ have not specifically examined the prognostic value of neuroimaging (such as lesion location^9^ and corticospinal tract (CST) damage using fractional anisotropy (FA)^10^) or neurophysiological biomarkers (such as lower limb motor evoked potential (MEP) status assessed using transcranial magnetic stimulation (TMS)^11^) measured within the first 14-days post-stroke. This overlooks the critical importance of the early window where key decisions regarding rehabilitation and discharge are made. An additional limitation is inclusion of cross-sectional designs which limit examination the prognostic utility of biomarkers to understand longer term mobility outcomes. This systematic review addresses these gaps by synthesizing only longitudinal studies that collected an early neuroimaging or neurophysiological biomarkers to examine mobility outcomes up to 24-months post-stroke. The findings from this review could advance clinical practice and research into mobility rehabilitation and recovery after stroke.

This systematic review aimed to examine the prognostic capacity of early neuroimaging and neurophysiological biomarkers of mobility outcomes up to 24-months post-stroke. For this review, mobility outcomes are defined as measures of impairment or activity limitations related to mobility. Impairments refer to lower limb body structure and function such as strength, while activity limitations refer to the ability to transfer or walk^12^. The review considers both the stage of prognostic development, based on the statistical approach, and the prognostic capacity of biomarkers, to distinguish between those that require further development (i.e., promising associative, discrimination/classification capacity), and those that may be considered ready for clinical implementation (i.e., promising validated predictive capacity).

## Methods

This systematic review was prospectively registered with the International Prospective Register of Systematic Reviews (ID CRD42022350771). The search was completed to identify both mobility and upper limb studies. We only report on papers with a mobility outcome in this review due to the large volume of studies identified and heterogenous findings. The Preferred Reporting Items for Systematic Reviews and Meta-Analysis (PRISMA) 2020 statement provided the framework for reporting^13^.

### Information Sources

Electronic searches of Ovid MEDLINE and EMBASE databases were conducted on 16 August 2022 and updated on 4 June 2025. Conference abstracts were linked to published reports where possible. Reference lists of included studies and other systematic reviews identified during title and abstract screening were hand searched to identify additional studies not identified in the search strategy.

### Search Strategy

Medical Subject Heading terms and keywords pertaining to stroke AND neuroimaging OR neurophysiological biomarkers AND mobility were used (Table S1). The Ingui Filter^14^ and keywords related to association were used to capture studies evaluating biomarker prognostic capacity. The Ingui Filter is recommended by the Cochrane Prognosis Methods Group^15^ as a search filter for prognostic and prediction studies. Consultation with experts in stroke informed preliminary searches before running the final search. There was no restriction based on publication date or language.

### Eligibility Criteria

Cohort studies enrolling human adult (≥18 years of age) stroke survivors who underwent early neuroimaging or neurophysiological biomarker collection (≤14-days post-stroke), measured mobility outcomes (defined as impairments of lower limb body structure or function such as strength, or activity limitations such as the ability to transfer or walk), both early (≤14-days post-stroke) and at ≥1 future time (>14-days and ≤24-months post-stroke) were included. Studies were required to report biomarker prognostic capacity results (association, discrimination/classification, or validation analyses) for at least one neuroimaging or neurophysiological biomarker in relation to mobility outcomes >14-days and ≤24-months post-stroke. The 14-day time window for biomarker collection was selected to extend beyond the SRRR-defined acute phase (≤7-days post-stroke)^16^ to ensure we captured more severe cohorts that may have included patients who were medically unstable or in intensive care units during the initial days post-stroke. All other study designs (e.g., clinical trials, cross sectional, systematic reviews), animal populations or humans with other neurological conditions (e.g., Parkinson’s Disease), neuroimaging or neurophysiological biomarkers collected during the sub-acute or chronic phase post-stroke (>14-days post-stroke), or outcome measures not specifically related to mobility (e.g., National Institutes of Health Stroke Scale (NIHSS) total score, modified Rankin scale (mRS)) were excluded.

### Selection Process

All identified studies were imported into Covidence and duplicates were removed. Titles and abstracts were independently screened by two reviewers (CL and JKF/KSH/ED). Full text for potentially relevant studies were retrieved and independently reviewed by two reviewers (CL and JKF/KSH/ED). Disagreements were resolved by consensus or discussion with a third independent reviewer (JKF/KSH/ED). Corresponding authors were contacted where there was uncertainty regarding eligibility for inclusion (maximum of two emails sent).

### Data Extraction Process

A custom-built Excel spreadsheet was developed for data collection in collaboration with a statistician (LC). Data were extracted independently by one author (CL) and cross checked by a second author (KSH/ED). Any disagreements were resolved by consensus or through discussion with a third reviewer as required (KSH/ED). All statistical analysis queries were discussed with a statistician (LC). Corresponding authors were contacted to obtain missing data (maximum of two emails sent). Non-English papers were translated by individuals who were fluent in the article language. Separate reports deemed to potentially represent the same study sample, were confirmed based on author name(s), study location, date and duration and participant demographics in alignment with Cochrane guidance^17^. Corresponding authors were contacted if uncertainty remained. When multiple reports were confirmed to relate to the same study sample, data were collated and participant data extracted for the primary report only.

### Data Extraction Items

Data were extracted for each study into the spreadsheet across the following categories:

1. Study information: Sample size and country of publication, eligibility criteria related to first stroke, cognition, communication and baseline mobility (impairment or activity).
2. Study participants: Mean/median study sample age, proportion of male and female participants, mean/median baseline stroke severity (defined by e.g., NIHSS), mean/median baseline mobility severity (impairment or activity), proportion of ischemic and hemorrhagic participants and proportion of participants that received medical intervention (e.g., thrombolysis).
3. Report biomarker(s): Biomarkers used in the analysis were classified as neuroimaging or neurophysiological measures, and their modality, metric and anatomical region (inclusive of region, tract, muscle or nerve) of interest were extracted. A maximum of three anatomical regions of interest per metric were extracted. Selection was prioritized based on their relevance to motor recovery (e.g., primary motor cortex over putamen, corona radiata over thalamus). The mean/median time post-stroke that the biomarkers were measured was also recorded and transformed into days (assuming 30-days per month).
4. Report outcome measure(s): Mobility outcomes were classified as impairment (lower limb body structure or function such as strength) or activity measures (ability to transfer or walk), consistent with the World Health Organization International Classification of Functioning, Disability and Health framework^12^ and the international SRRR taskforce recommendations^16^. Mean/median time post-stroke that the outcomes were measured were recorded, transformed into months and compared to determine similarities between studies. If these data were not available, studies were classified based on their methods. The follow-up timepoints of interest were 1-, 3-, 6- and 12-months post-stroke, and these were allocated to a post-stroke recovery epoch. The SRRR taskforce defined recovery epochs were used: early subacute, >7-days but ≤3-months; late subacute, >3-months but ≤6-months; chronic, >6-months^18^.
5. Report biomarker prognostic capacity: Method of statistical analysis used to determine biomarker prognostic capacity (e.g., Spearman’s correlation, linear regression, receiver operating characteristic (ROC) analysis) and results from the analysis (i.e., magnitude and/or numerical values of significance (i.e., *p* values)) where provided. All data were extracted as reported; we did not correct for any inconsistencies.

### Data Synthesis

In line with Cochrane guidance^17^, meta-analyses could not be performed due to heterogeneity in biomarkers, outcome measures, follow-up time points and methods of statistical analyses performed across reports. Findings were summarized descriptively and evaluated at the level of individual biomarker analyses. Summary tables were organized by biomarker type (neuroimaging or neurophysiological) and follow-up recovery epoch, with mobility outcomes presented separately for impairment and activity limitation measures.

Each biomarker analysis was classified into one of three categories based on the type of statistical analysis approach used, irrespective of the terminology reported by individual studies: 1) association, 2) discrimination/classification or 3) validation (Table 1a). The magnitude of relevant statistical measures was used as the primary indicator for interpreting prognostic capacity (Table 1b). Magnitude ratings were applied where appropriate, depending on the type of statistical measure reported. Correlation coefficients were rated according to established statistical standards^19^. As no universally accepted or guideline recommended thresholds^20,21^ exist for area under the curve (AUC), sensitivity or specificity, magnitude ratings were informed by published literature^22^ and authorship group consensus. Linear and logistic regression coefficients were not rated for prognostic capacity based on magnitude, as their interpretation depends on the scale and units of the biomarker and outcome measures, limiting comparability across studies. Where these measures were reported, or where no magnitude was provided, statistical significance (*p* values) despite acknowledged limitations^23^ was used as the main indicator of prognostic capacity. Statistical significance was rated according to numerical *p* values of less than or equal to 0.05 (rated significant) or greater than 0.05 (rated non-significant). Accuracy and positive/negative predictive values were not rated for prognostic capacity, due to their dependence on outcome prevalence, which restricts generalizability to broader populations.

**Table 1a.** Illustrative Examples of Statistical Analysis Approach.

| Statistical analysis approach | Example methods of statistical analysis |
| --- | --- |
| Association | Pearson correlation, Spearman's correlation, linear regression, logistic regression |
| Discrimination/classification | Receiver operating characteristic analysis, sensitivity, specificity |
| Validation | In addition to an appropriate statistical analysis, internal validation (e.g., random split of data, bootstrap) or external validation (e.g., temporal, geographical) |

**Table 1b.** Prognostic Magnitude Ratings.

| Prognostic statistical measure | Magnitude rating |
| --- | --- |
| **Association** |  |
| Pearson correlation coefficient<sup>19</sup> | Small: 0.1-0.3<br>Moderate: 0.3-0.5<br>Large: >0.5 |
| Spearman's correlation coefficient<sup>19</sup> | Small: 0.1-0.3<br>Moderate: 0.3-0.5<br>Large: >0.5 |
| Linear and logistic regression coefficient | Not rated by magnitude; prognostic capacity interpreted using statistical significance threshold ($p \leq 0.05$). |
| **Discrimination/classification** |  |
| Area under the curve<sup>22</sup> | Poor: <0.5<br>Fair: 0.5-0.8<br>Excellent: >0.8 |
| Sensitivity or specificity | Poor: <50%<br>Fair: 50-80%<br>Excellent: >80% |
| Accuracy or positive/negative predictive | Not rated |
| values |  |

### Risk of Bias Assessment

Risk of bias was assessed at the level of report. In accordance with the Cochrane Prognosis Methods Group^15^, risk of bias was assessed using the Quality in Prognostic Factor Studies (QUIPS) tool across six domains^24^. Confounding factors considered relevant for this review were prespecified as age^1^ and baseline stroke severity^1^. Following QUIPS guidelines, an overall summary risk of bias judgement for each report was not defined. One reviewer (CL) independently rated each report. A second reviewer (BCVC/ED) independently cross-checked 20% of reports. Any discrepancies or queries were resolved by consensus between reviewers or through discussion with a third reviewer (LC) if required.

### Data Availability Statement

The data that support the findings of this systematic review are available from the corresponding author upon reasonable request.

## Results

### Study Selection

The search strategy yielded 37,191 records (MEDLINE n=8,828, Embase n=28,363) with 26,486 records remaining after duplicates were removed. A total of 459 full text records were assessed for eligibility. Twenty reports fulfilled all eligibility criteria, which refer to 18 independent study samples. The most common reasons for exclusion at full text were wrong outcome measure (n=152) and wrong biomarker measurement timepoint (n=115). Six studies were excluded as authors did not respond to confirm the initial biomarker measurement timepoint. From review of included study references, two additional studies were identified that fulfilled the inclusion criteria^25,26^. The PRISMA flowchart for the identification and inclusion of studies is shown in Figure 1.

**Figure 1.**
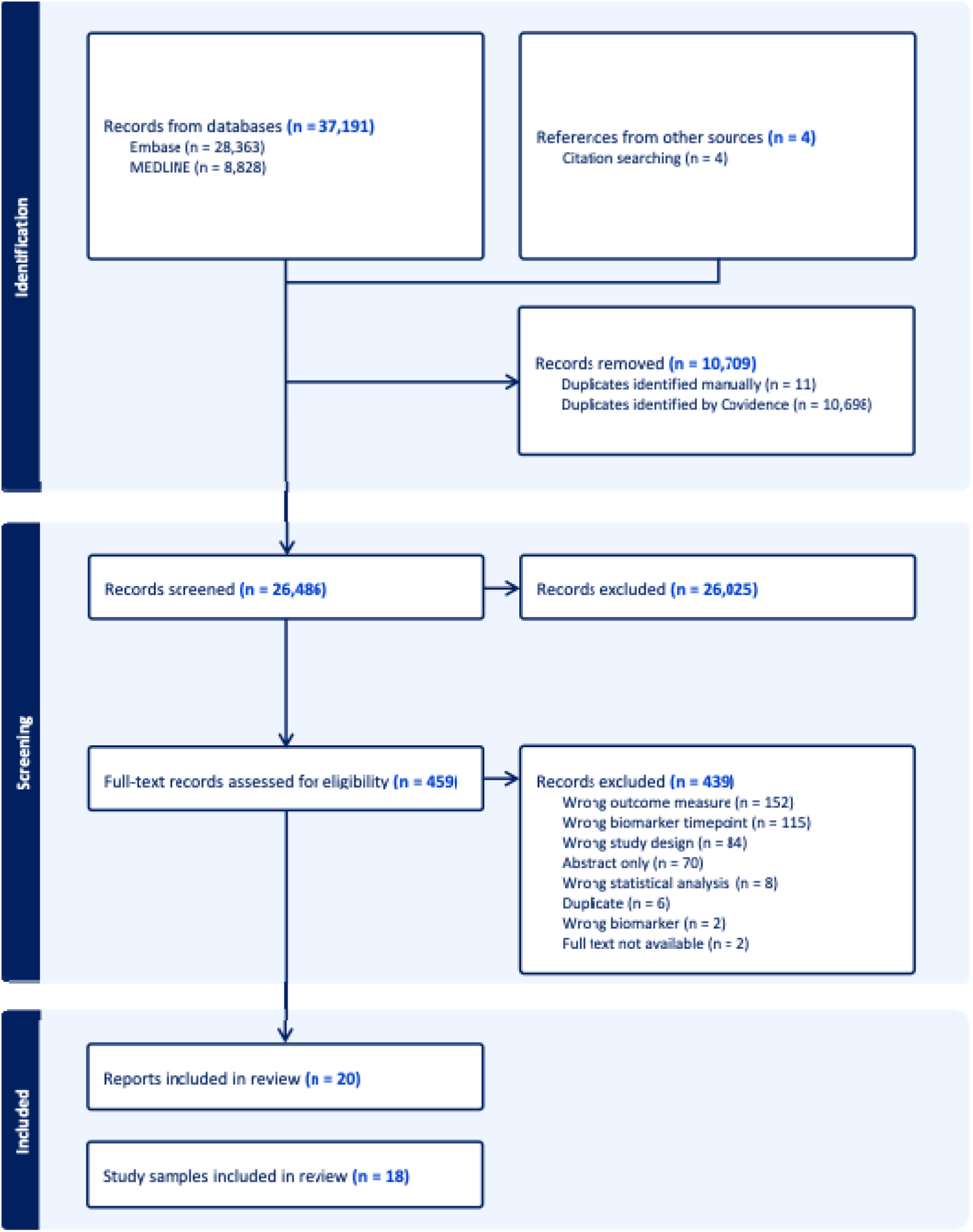
PRISMA Flowchart.

### Study Characteristics

Table 2 presents study characteristics. A total of 2,160 participants were included across 18 studies. The median (range) sample size was 40 (17-1233). Most studies were conducted in Asia (61%). The included study samples reflected primarily people who had a first (67% of studies included an eligibility criterion) ischemic (72%) stroke with no cognitive (44%) or communication deficits (33%). The mean (SD) age of participants across studies was 62 (5) years and there were similar proportions of males (55%) and females (45%) enrolled. Due to insufficient reporting (only 4/18 studies), acute medical intervention, baseline stroke severity and baseline mobility severity could not be summarized. Of the 18 studies, 15 investigated neuroimaging biomarkers and six investigated neurophysiological biomarkers. The most common follow-up time point was 3-months (early subacute, n=9), followed by 6-months (late-subacute, n=8) post-stroke. Eleven studies measured impairments and 12 measured activity limitations. Nine authors were contacted to provide additional biomarker prognostic capacity results, of which two returned additional data.

**Table 2.**
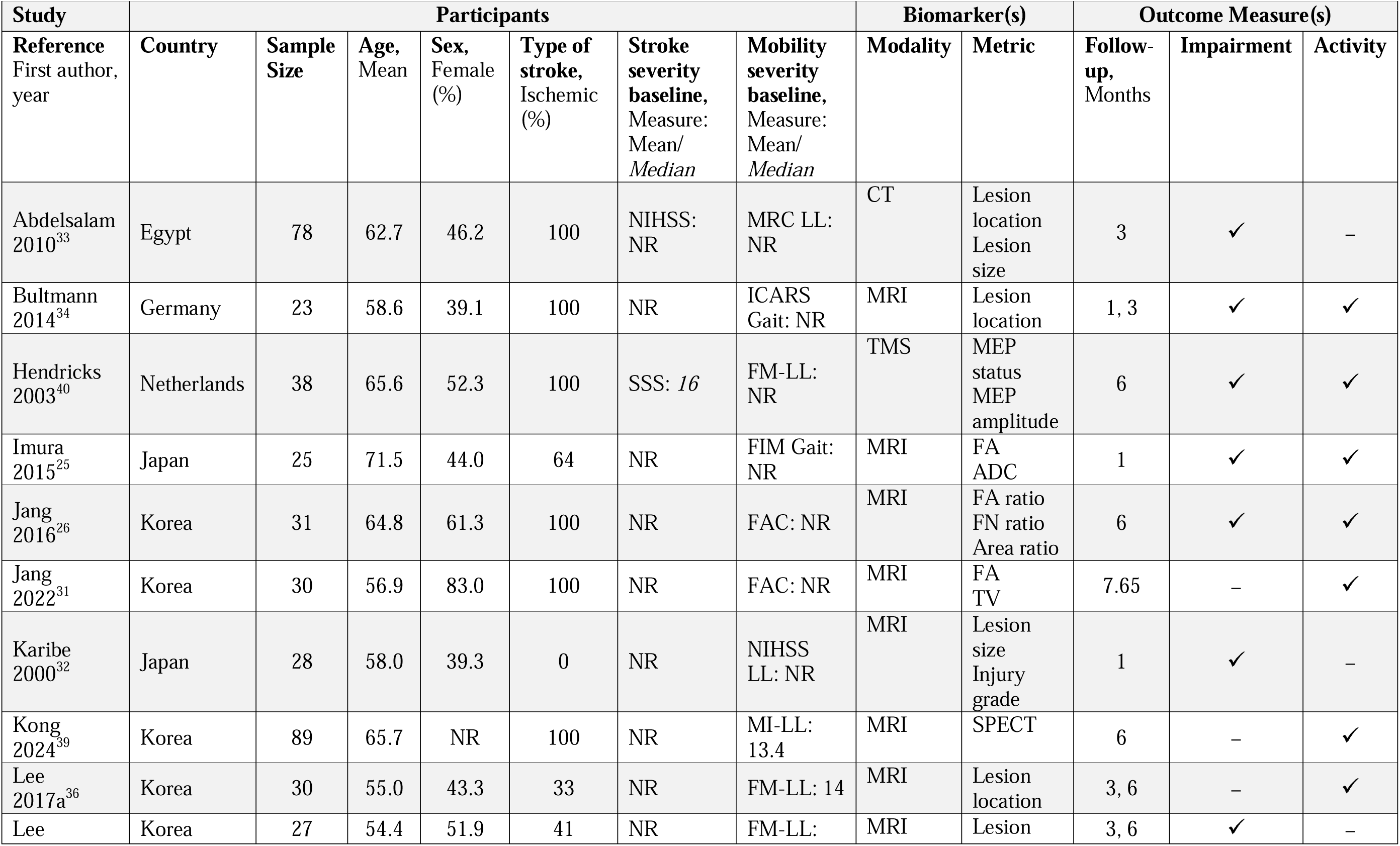
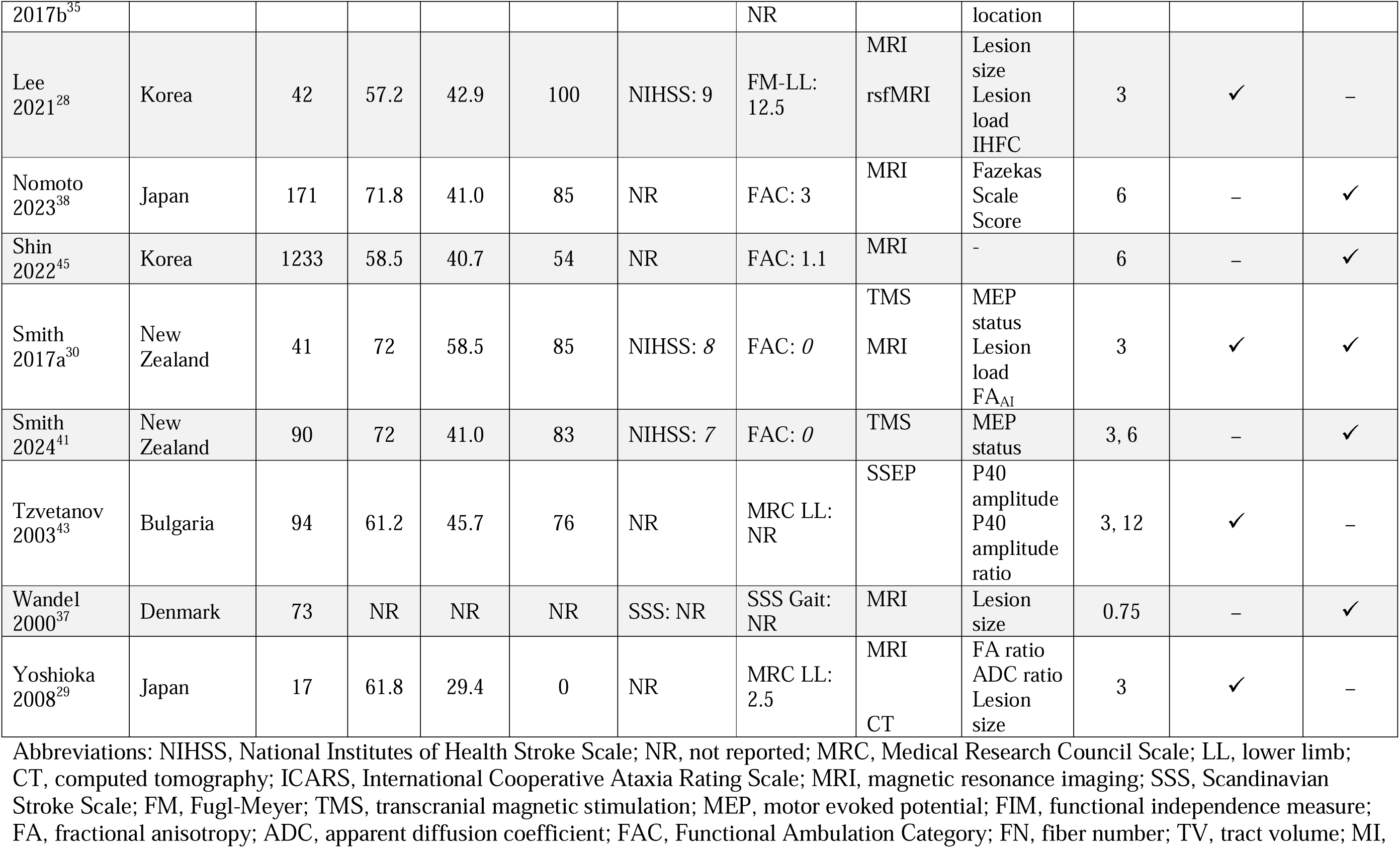
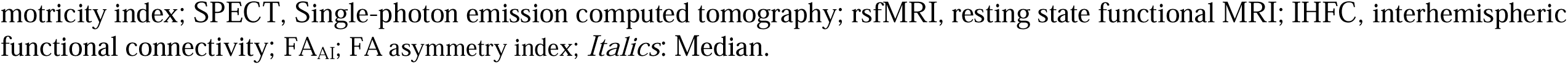
Study Characteristics.

| Study | Participants |  |  |  |  |  |  | Biomarker(s) |  | Outcome Measure(s) |  |  |
| --- | --- | --- | --- | --- | --- | --- | --- | --- | --- | --- | --- | --- |
| Reference<br>First author,<br>year | Country | Sample<br>Size | Age,<br>Mean | Sex,<br>Female<br>(%) | Type of<br>stroke,<br>Ischemic<br>(%) | Stroke<br>severity<br>baseline,<br>Measure:<br>Mean/<br>Median | Mobility<br>severity<br>baseline,<br>Measure:<br>Mean/<br>Median | Modality | Metric | Follow-up,<br>Months | Impairment | Activity |
| Abdelsalam<br>2010<sup>33</sup> | Egypt | 78 | 62.7 | 46.2 | 100 | NIHSS:<br>NR | MRC LL:<br>NR | CT | Lesion<br>location<br>Lesion<br>size | 3 | ✓ | – |
| Bultmann<br>2014<sup>34</sup> | Germany | 23 | 58.6 | 39.1 | 100 | NR | ICARS<br>Gait: NR | MRI | Lesion<br>location | 1, 3 | ✓ | ✓ |
| Hendricks<br>2003<sup>40</sup> | Netherlands | 38 | 65.6 | 52.3 | 100 | SSS: 16 | FM-LL:<br>NR | TMS | MEP<br>status<br>MEP<br>amplitude | 6 | ✓ | ✓ |
| Imura<br>2015<sup>25</sup> | Japan | 25 | 71.5 | 44.0 | 64 | NR | FIM Gait:<br>NR | MRI | FA<br>ADC | 1 | ✓ | ✓ |
| Jang<br>2016<sup>26</sup> | Korea | 31 | 64.8 | 61.3 | 100 | NR | FAC: NR | MRI | FA ratio<br>FN ratio<br>Area ratio | 6 | ✓ | ✓ |
| Jang<br>2022<sup>31</sup> | Korea | 30 | 56.9 | 83.0 | 100 | NR | FAC: NR | MRI | FA<br>TV | 7.65 | – | ✓ |
| Karibe<br>2000<sup>32</sup> | Japan | 28 | 58.0 | 39.3 | 0 | NR | NIHSS<br>LL: NR | MRI | Lesion<br>size<br>Injury<br>grade | 1 | ✓ | – |
| Kong<br>2024<sup>39</sup> | Korea | 89 | 65.7 | NR | 100 | NR | MI-LL:<br>13.4 | MRI | SPECT | 6 | – | ✓ |
| Lee<br>2017a<sup>36</sup> | Korea | 30 | 55.0 | 43.3 | 33 | NR | FM-LL: 14 | MRI | Lesion<br>location | 3, 6 | – | ✓ |
| Lee | Korea | 27 | 54.4 | 51.9 | 41 | NR | FM-LL: | MRI | Lesion | 3, 6 | ✓ | – |
| 2017b<sup>35</sup> |  |  |  |  |  |  | NR |  | location |  |  |  |
| Lee 2021<sup>28</sup> | Korea | 42 | 57.2 | 42.9 | 100 | NIHSS: 9 | FM-LL: 12.5 | MRI<br>rsfMRI | Lesion size<br>Lesion load<br>IHFC | 3 | ✓ | – |
| Nomoto 2023<sup>38</sup> | Japan | 171 | 71.8 | 41.0 | 85 | NR | FAC: 3 | MRI | Fazekas Scale Score | 6 | – | ✓ |
| Shin 2022<sup>45</sup> | Korea | 1233 | 58.5 | 40.7 | 54 | NR | FAC: 1.1 | MRI | - | 6 | – | ✓ |
| Smith 2017a<sup>30</sup> | New Zealand | 41 | 72 | 58.5 | 85 | NIHSS: 8 | FAC: 0 | TMS<br>MRI | MEP status<br>Lesion load<br>FA<sub>AI</sub> | 3 | ✓ | ✓ |
| Smith 2024<sup>41</sup> | New Zealand | 90 | 72 | 41.0 | 83 | NIHSS: 7 | FAC: 0 | TMS | MEP status | 3, 6 | – | ✓ |
| Tzvetanov 2003<sup>43</sup> | Bulgaria | 94 | 61.2 | 45.7 | 76 | NR | MRC LL: NR | SSEP | P40 amplitude<br>P40 amplitude ratio | 3, 12 | ✓ | – |
| Wandel 2000<sup>37</sup> | Denmark | 73 | NR | NR | NR | SSS: NR | SSS Gait: NR | MRI | Lesion size | 0.75 | – | ✓ |
| Yoshioka 2008<sup>29</sup> | Japan | 17 | 61.8 | 29.4 | 0 | NR | MRC LL: 2.5 | MRI<br>CT | FA ratio<br>ADC ratio<br>Lesion size | 3 | ✓ | – |

Abbreviations: NIHSS, National Institutes of Health Stroke Scale; NR, not reported; MRC, Medical Research Council Scale; LL, lower limb; CT, computed tomography; ICARS, International Cooperative Ataxia Rating Scale; MRI, magnetic resonance imaging; SSS, Scandinavian Stroke Scale; FM, Fugl-Meyer; TMS, transcranial magnetic stimulation; MEP, motor evoked potential; FIM, functional independence measure; FA, fractional anisotropy; ADC, apparent diffusion coefficient; FAC, Functional Ambulation Category; FN, fiber number; TV, tract volume; MI,
motricity index; SPECT, Single-photon emission computed tomography; rsfMRI, resting state functional MRI; IHFC, interhemispheric functional connectivity; FA<sub>AI</sub>, FA asymmetry index; *Italics*: Median.

### Risk of Bias Assessment

Table 3 presents QUIPS tool ratings. The domains of *study confounding* (n=17, 85%), *study participation* (n=15, 75%) and *statistical analysis and reporting* (n=13, 65%), had the highest risk of bias across reports. Most reports were rated as moderate risk for *study confounding* as they did not account for age and baseline stroke severity in study design or analyses. However, it is important to note that confounding bias is not typically applicable to associative studies, where causal inference is not intended^27^. *Study participation* was rated as high risk of bias in five (25%) reports and moderate in ten (50%) reports, primarily due to insufficient detail describing the baseline sample and recruitment procedures. The *statistical analysis and reporting* domain was rated as high risk of bias in twelve (60%) reports and moderate in one (5%) report, largely due to selective reporting of results. In contrast, all reports used *objective measures of mobility impairment and activity limitation* and were judged as low risk of bias. Eighteen (90%) reports were rated as low risk of bias for *prognostic factor measurement*, with two (10%) studies as moderate due to limited description of how the biomarker was measured.

**Table 3.** QUIPS Risk of Bias Assessment.

| Reference<br>First author, year | Study<br>Participation | Study Attrition | Prognostic Factor<br>Measurement | Outcome<br>Measurement | Study<br>Confounding | Statistical Analysis<br>and Reporting |
| --- | --- | --- | --- | --- | --- | --- |
| Abdelsalam 2010<sup>33</sup> | High | High | Moderate | Low | Moderate | High |
| Bultmann 2014<sup>34</sup> | High | Moderate | Moderate | Low | Moderate | High |
| Hendricks 2003<sup>40</sup> | Low | Low | Low | Low | Moderate | High |
| Imura 2015<sup>25</sup> | High | High | Low | Low | Moderate | High |
| Jang 2016<sup>26</sup> | Moderate | High | Low | Low | Moderate | Low |
| Jang 2022<sup>31</sup> | Moderate | High | Low | Low | Moderate | Low |
| Karibe 2000<sup>32</sup> | Moderate | High | Low | Low | Moderate | Moderate |
| Kong 2024<sup>39</sup> | Low | Moderate | Low | Low | Low | High |
| Lee 2017a<sup>36</sup> | Moderate | Low | Low | Low | Moderate | High |
| Lee 2017b<sup>35</sup> | Moderate | Low | Low | Low | Moderate | High |
| Lee 2021<sup>28</sup> | Moderate | High | Low | Low | Moderate | High |
| Nomoto 2023<sup>38</sup> | Low | Low | Low | Low | Low | Low |
| Shin 2022<sup>45</sup> | Low | High | Low | Low | Moderate | Low |
| Smith 2017a<sup>30</sup> | Moderate | Low | Low | Low | Moderate | High |
| Smith 2017b<sup>42</sup> | Moderate | Low | Low | Low | Moderate | Low |
| Smith 2024<sup>41</sup> | Moderate | Low | Low | Low | Low | High |
| Tzvetanov 2003<sup>43</sup> | High | Low | Low | Low | Moderate | Low |
| Tzvetanov 2005<sup>44</sup> | High | Low | Low | Low | Moderate | Low |
| Wandel 2000<sup>37</sup> | Low | Low | Low | Low | Moderate | High |
| Yoshioka 2008<sup>29</sup> | Moderate | Low | Low | Low | Moderate | High |

Abbreviations: QUIPS, Quality in Prognostic Factor Studies
Rating: Red = high, yellow = medium, green = low

### Biomarkers

The median time to initial biomarker measurement was 7.5 (range 1.8-14) days. Eighty-six biomarker analyses were reported, including 61 neuroimaging and 25 neurophysiological analyses. Most magnetic resonance imaging (MRI) biomarkers were collected using a 1.5T scanner (71.4%).

### Outcome Measures

Six different mobility activity measures were used across 50 analyses; functional ambulation categorization (FAC, n=41 analyses) was the most common. Five mobility impairment measures were used across 36 analyses; Fugl Meyer lower limb (FM LL, n=15 analyses) was the most common. Activity measures were predominantly assessed in the late subacute phase (n=24 analyses), while impairment measures were more often reported in the early subacute phase (n=26 analyses)

### Biomarker Prognostic Capacity

Magnitude of prognostic statistical measures was reported for 59.3% (n=51) of biomarker analyses. Statistical significance was reported for most analyses (n=70, 81.4%), while six (6.9%) did not report any measure of prognostic capacity. Tables 4a to 4f present the findings for magnitude and significance values by biomarker metric.

**Table 4a.** Early Subacute: Neuroimaging Biomarker Performance.

| Reference<br>First author,<br>year, country | Sample<br>Size | Outcome<br>measure | Neuroimaging Biomarker |  |  | Method of<br>Statistical Analysis | Statistical<br>Analysis<br>Category | Prognostic Capacity |  |
| --- | --- | --- | --- | --- | --- | --- | --- | --- | --- |
|  |  |  | Modality | Metric | Region / Tract of<br>Interest |  |  | Magnitude | Significance |
| Impairment, n = 22 analyses |  |  |  |  |  |  |  |  |  |
| Abdelsalam<br>2010<br>Egypt<sup>33</sup> | 78 | MRC LL | CT | *Lesion*<br>*location* | Cortical | Logistic regression<br>(Coefficient,<br>95% CI) | Association | NR | NR |
|  | 78 | MRC LL | CT | *Lesion*<br>*location* | Subcortical | Logistic regression<br>(Coefficient,<br>95% CI) | Association | 0.016 (0.000-0.671) | p = 0.03 |
|  | 78 | MRC LL | CT | *Lesion*<br>*location* | Combined cortical<br>and subcortical | Logistic regression<br>(Coefficient,<br>95% CI) | Association | NR | NR |
|  | 78 | MRC LL | CT | *Lesion*<br>*size* | - | Logistic regression<br>(Coefficient,<br>95% CI) | Association | NR | NR |
| Bultmann<br>2014<br>Germany<sup>34</sup> | 16 | ICARS LL | T2 | *Lesion*<br>*location* | SCA / PICA<br>territory | ANOVA | Association | - | p > 0.08 |
|  | 16 | ICARS LL | T2 | *Lesion*<br>*location* | Cerebellar nuclei<br>involvement | ANOVA | Association | - | p > 0.8 |
| Imura<br>2015<br>Japan<sup>25</sup> | 25 | MI LL | DTI | FA | Corticospinal tract | Spearman’s<br>correlation (r<sub>s</sub>) | Association | 0.60 | p < 0.05 |
|  | 25 | MI LL | DTI | ADC | Corticospinal tract | Spearman’s<br>correlation (r<sub>s</sub>) | Association | 0.39 | p = 0.06 |
| Karibe<br>2000<br>Japan<sup>32</sup> | 28 | NIHSS LL | DWI | *Injury*<br>*grade* | Corticospinal tract | Spearman’s<br>correlation (r<sub>s</sub>) | Association | 0.868 | p < 0.001 |
|  | 28 | NIHSS LL | T2, DWI | Lesion<br>size | - | Spearman’s<br>correlation (r<sub>s</sub>) | Association | 0.663 | NR |
| Lee<br>2017b | 27 | *FM LL* | T1, T2 | *Lesion*<br>*location* | Motor cortex | Fishers exact test | Association | - | p > 0.05 |
| Korea<sup>35</sup> | 27 | FM LL | T1, T2 | Lesion location | Posterior half middle third corona radiata | Fishers exact test | Association | - | p > 0.05 |
|  | 27 | FM LL | T1, T2 | Lesion location | Posterior limb internal capsule | Fishers exact test | Association | - | p > 0.05 |
| Lee 2021 Korea<sup>28</sup> | 42 | FM LL | DTI | Lesion load | Corticospinal tract | Linear regression (Coefficient, 95% CI) | Association | NR | p = 0.319 |
|  | 42 | FM LL | T1 | Lesion size | - | Linear regression (Coefficient, 95% CI) | Association | NR | p = 0.383 |
|  | 42 | FM LL | DTI | FA | Ipsilesional corticospinal tract | Linear regression (Coefficient, 95% CI) | Association | NR | p = 0.004 |
|  | 42 | FM LL | DTI | FA | Contralesional corticospinal tract | Linear regression (Coefficient, 95% CI) | Association | NR | p = 0.026 |
|  | 42 | FM LL | rsfMRI | IHFC | - | Linear regression (Coefficient, 95% CI) | Association | NR | p = 0.323 |
| Smith 2017b New Zealand<sup>42</sup> | 23 | FM LL | T1, DWI | Lesion load | Corticospinal tract | Linear regression (Coefficient, 95% CI) | Association | 0.051 (0.056-0.158) | p = 0.33 |
| Yoshioka 2008 Japan<sup>29</sup> | 17 | MRC LL | DTI | FA ratio | Corticospinal tract | Spearman's correlation (r<sub>s</sub>) | Association | 0.56 | p < 0.05 |
|  | 17 | MRC LL | DTI | ADC ratio | Corticospinal tract | Spearman's correlation (r<sub>s</sub>) | Association | NR | p > 0.05 |
|  | 17 | MRC LL | CT | Lesion size | NA | Spearman's correlation (r<sub>s</sub>) | Association | NR | p > 0.05 |
| Activity limitation, n = 13 analyses |  |  |  |  |  |  |  |  |  |
| Bultmann 2014 Germany<sup>34</sup> | 16 | ICARS Gait | T2 | Lesion location | SCA / PICA territory | ANOVA | Association | - | p > 0.08 |
|  | 16 | ICARS Gait | T2 | Lesion location | Cerebellar nuclei involvement | ANOVA | Association | - | p > 0.8 |
| Imura<br>2015<br>Japan<sup>25</sup> | 25 | FIM Gait | DTI | FA | Corticospinal tract | Spearman's correlation ($r_s$) | Association | 0.67 | p < 0.05 |
| | 25 | FIM Gait | DTI | ADC | Corticospinal tract | Spearman's correlation ($r_s$) | Association | 0.19 | p = 0.36 |
| | 25 | BI Gait | DTI | FA | Corticospinal tract | Spearman's correlation ($r_s$) | Association | 0.65 | p < 0.05 |
| | 25 | BI Gait | DTI | ADC | Corticospinal tract | Spearman's correlation ($r_s$) | Association | 0.21 | p = 0.30 |
| Lee<br>2017a<br>Korea<sup>36</sup> | 30 | *FAC* | T1, T2 | *Lesion location* | Primary motor cortex | Wilcoxon's signed-rank test | Association | - | p < 0.05 |
|  | 30 | *FAC* | T1, T2 | *Lesion location* | Corona radiata | Wilcoxon's signed-rank test | Association | - | p < 0.05 |
|  | 30 | *FAC* | T1, T2 | *Lesion location* | Internal capsule | Wilcoxon's signed-rank test | Association | - | p < 0.05 |
| Smith<br>2017a<br>New Zealand<sup>30</sup> | 30 | *FAC* | T1, DWI | Lesion load | Corticospinal tract | Logistic regression (Coefficient, 95% CI) | Association | 0.984 (0.937-1.034) | p = 0.527 |
|  | 30 | *FAC* | T1, DWI | Lesion load | Sensorimotor tract | Logistic regression (Coefficient, 95% CI) | Association | 0.983 (0.926-1.044) | p = 0.586 |
|  | 30 | *FAC* | T1, DWI | FA<sub>AI</sub> | Corticospinal tract | Logistic regression (Coefficient, 95% CI) | Association | 0.372 (0.002-83.709) | p = 0.72 |
| Wandel<br>2000<br>Denmark<sup>37</sup> | 73 | *SSS Gait* | CT | Lesion size | - | Student t test | Association | - | p > 0.05 |

Abbreviations: MRC, Medical Research Council Scale; LL, lower limb; CT, computed tomography; CI, confidence interval; NR, not reported; ICARS, International Cooperative Ataxia Rating Scale; T2, T2-weighted imaging; SCA, superior cerebellar artery; PICA, posterior inferior cerebellar artery; ANOVA, analysis of variance; MI, motricity index; DTI, diffusion tensor imaging; FA, fractional anisotropy; $r_s$ , Spearman's correlation coefficient; ADC, apparent diffusion coefficient; NIHSS, National Institutes of Health Stroke Scale; DWI, diffusion-weighted imaging; FM, Fugl-Meyer Assessment; T1, T1-weighted imaging; rsfMRI, resting state functional magnetic resonance imaging (MRI); IHFC, interhemispheric homotopic functional connectivity; FIM, functional independence measure; BI, barthel index; SSS, Scandinavian Stroke Scale; FAC, functional ambulation category; FA<sub>AI</sub>, FA asymmetry index; *Italics* = Biomarker metric or outcome measure dichotomized or categorized; magnitude, association: red = small, yellow = moderate, green = large; significance: red = non-significant, green = significant.

**Table 4b.**
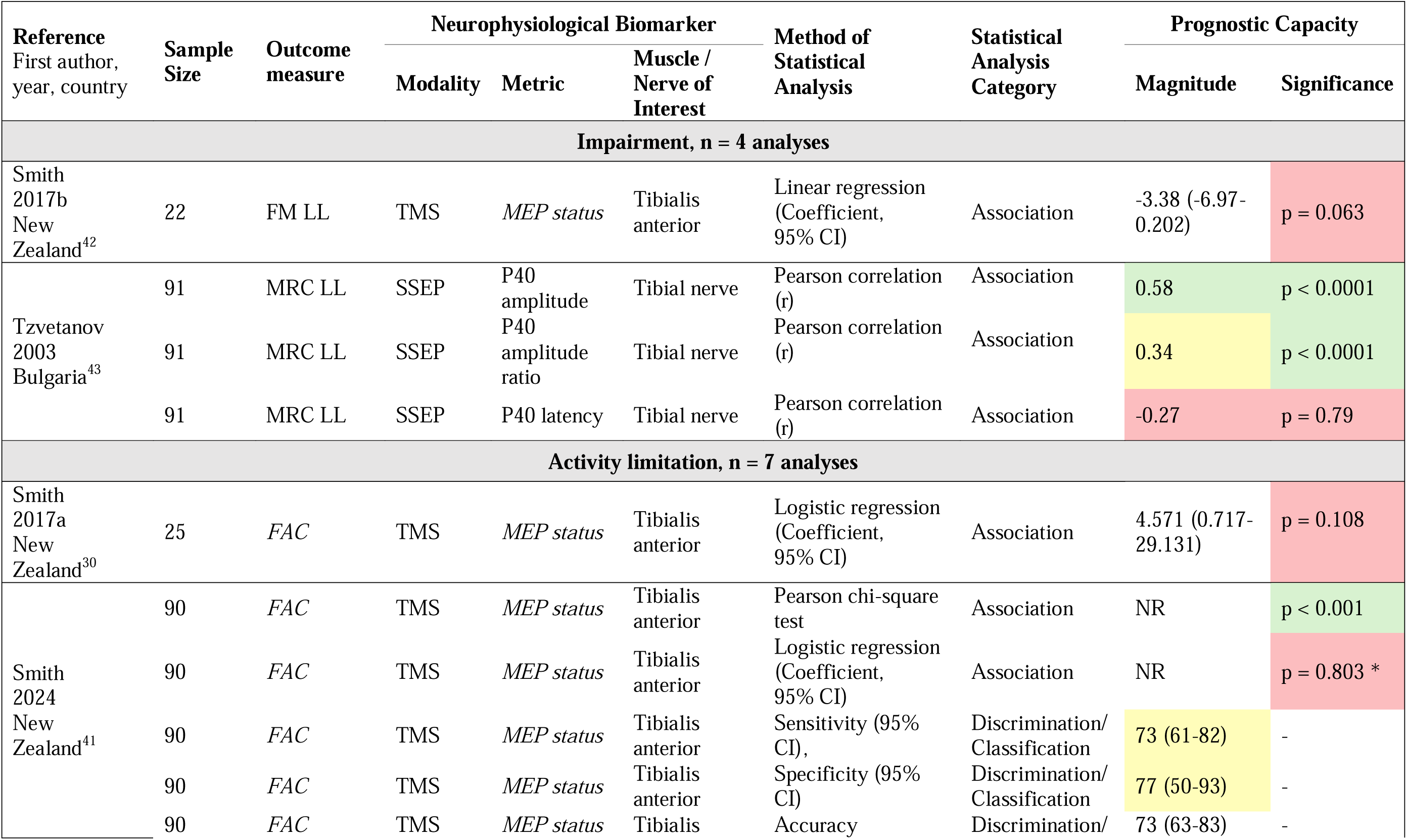
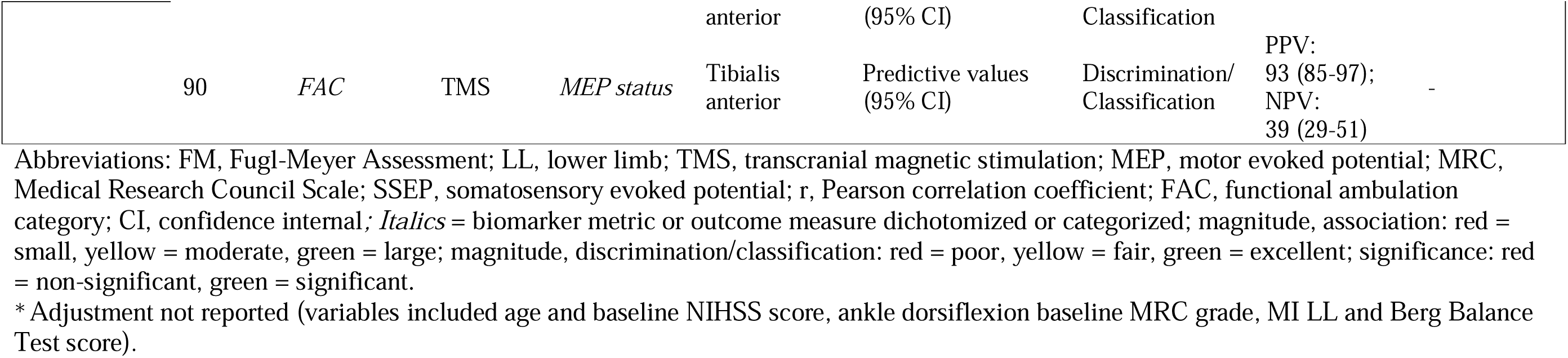
Early Subacute: Neurophysiological Biomarker Performance.

**Table 4c.**
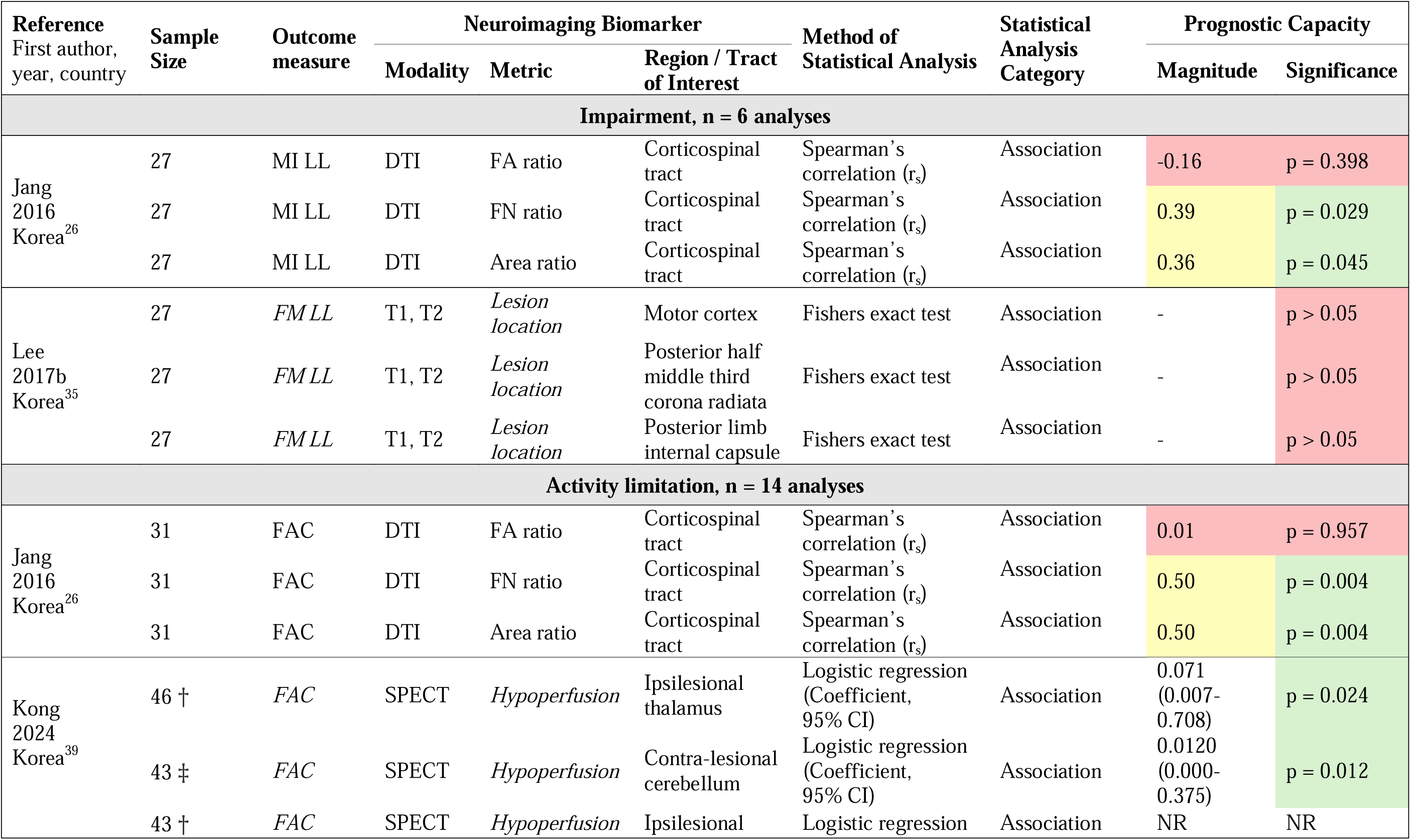
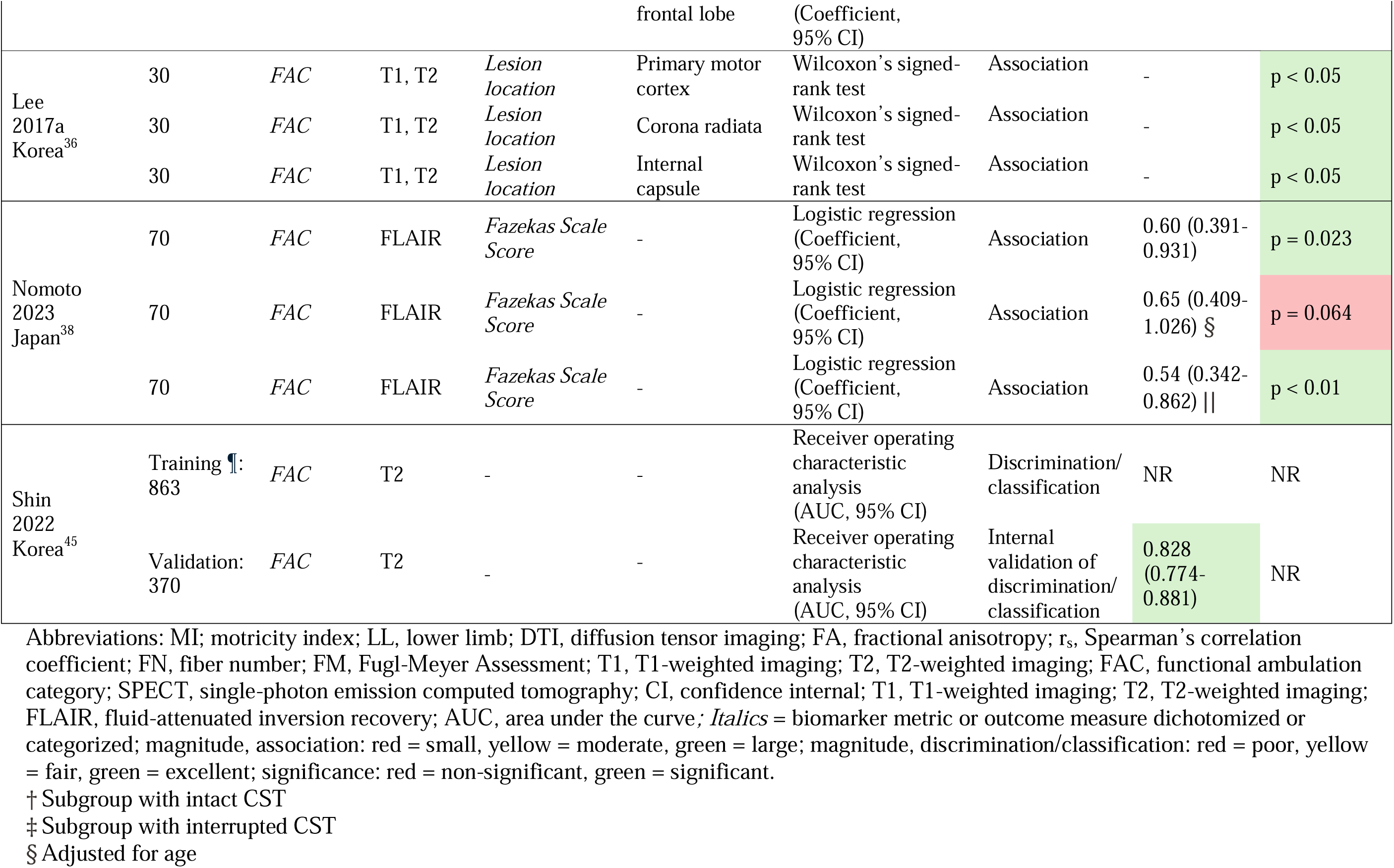
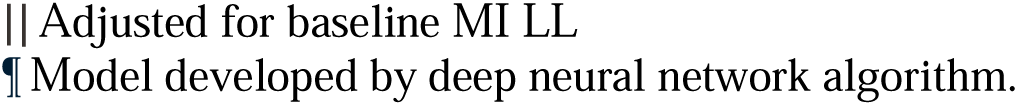
Late Subacute: Neuroimaging Biomarker Performance.

| Reference<br>First author,<br>year, country | Sample<br>Size | Outcome<br>measure | Neuroimaging Biomarker |  |  | Method of<br>Statistical Analysis | Statistical<br>Analysis<br>Category | Prognostic Capacity |  |
| --- | --- | --- | --- | --- | --- | --- | --- | --- | --- |
|  |  |  | Modality | Metric | Region / Tract<br>of Interest |  |  | Magnitude | Significance |
| Impairment, n = 6 analyses |  |  |  |  |  |  |  |  |  |
| Jang<br>2016<br>Korea<sup>26</sup> | 27 | MI LL | DTI | FA ratio | Corticospinal tract | Spearman's correlation (r<sub>s</sub>) | Association | -0.16 | p = 0.398 |
|  | 27 | MI LL | DTI | FN ratio | Corticospinal tract | Spearman's correlation (r<sub>s</sub>) | Association | 0.39 | p = 0.029 |
|  | 27 | MI LL | DTI | Area ratio | Corticospinal tract | Spearman's correlation (r<sub>s</sub>) | Association | 0.36 | p = 0.045 |
| Lee<br>2017b<br>Korea<sup>35</sup> | 27 | FM LL | T1, T2 | Lesion location | Motor cortex | Fishers exact test | Association | - | p > 0.05 |
|  | 27 | FM LL | T1, T2 | Lesion location | Posterior half middle third corona radiata | Fishers exact test | Association | - | p > 0.05 |
|  | 27 | FM LL | T1, T2 | Lesion location | Posterior limb internal capsule | Fishers exact test | Association | - | p > 0.05 |
| Activity limitation, n = 14 analyses |  |  |  |  |  |  |  |  |  |
| Jang<br>2016<br>Korea<sup>26</sup> | 31 | FAC | DTI | FA ratio | Corticospinal tract | Spearman's correlation (r<sub>s</sub>) | Association | 0.01 | p = 0.957 |
|  | 31 | FAC | DTI | FN ratio | Corticospinal tract | Spearman's correlation (r<sub>s</sub>) | Association | 0.50 | p = 0.004 |
|  | 31 | FAC | DTI | Area ratio | Corticospinal tract | Spearman's correlation (r<sub>s</sub>) | Association | 0.50 | p = 0.004 |
| Kong<br>2024<br>Korea<sup>39</sup> | 46 † | FAC | SPECT | Hypoperfusion | Ipsilesional thalamus | Logistic regression (Coefficient, 95% CI) | Association | 0.071 (0.007-0.708) | p = 0.024 |
|  | 43 ‡ | FAC | SPECT | Hypoperfusion | Contra-lesional cerebellum | Logistic regression (Coefficient, 95% CI) | Association | 0.0120 (0.000-0.375) | p = 0.012 |
|  | 43 † | FAC | SPECT | Hypoperfusion | Ipsilesional | Logistic regression | Association | NR | NR |

|  |  |  |  |  | frontal lobe | (Coefficient, 95% CI) |  |  |  |
| --- | --- | --- | --- | --- | --- | --- | --- | --- | --- |
| Lee 2017a Korea<sup>36</sup> | 30 | FAC | T1, T2 | *Lesion location* | Primary motor cortex | Wilcoxon's signed-rank test | Association | - | p < 0.05 |
|  | 30 | FAC | T1, T2 | *Lesion location* | Corona radiata | Wilcoxon's signed-rank test | Association | - | p < 0.05 |
|  | 30 | FAC | T1, T2 | *Lesion location* | Internal capsule | Wilcoxon's signed-rank test | Association | - | p < 0.05 |
| Nomoto 2023 Japan<sup>38</sup> | 70 | FAC | FLAIR | *Fazekas Scale Score* | - | Logistic regression (Coefficient, 95% CI) | Association | 0.60 (0.391-0.931) | p = 0.023 |
|  | 70 | FAC | FLAIR | *Fazekas Scale Score* | - | Logistic regression (Coefficient, 95% CI) | Association | 0.65 (0.409-1.026) § | p = 0.064 |
|  | 70 | FAC | FLAIR | *Fazekas Scale Score* | - | Logistic regression (Coefficient, 95% CI) | Association | 0.54 (0.342-0.862) \|\| | p < 0.01 |
| Shin 2022 Korea<sup>45</sup> | Training ¶: 863 | FAC | T2 | - | - | Receiver operating characteristic analysis (AUC, 95% CI) | Discrimination/ classification | NR | NR |
|  | Validation: 370 | FAC | T2 | - | - | Receiver operating characteristic analysis (AUC, 95% CI) | Internal validation of discrimination/ classification | 0.828 (0.774-0.881) | NR |

Abbreviations: MI; motricity index; LL, lower limb; DTI, diffusion tensor imaging; FA, fractional anisotropy; $r_s$ , Spearman's correlation coefficient; FN, fiber number; FM, Fugl-Meyer Assessment; T1, T1-weighted imaging; T2, T2-weighted imaging; FAC, functional ambulation category; SPECT, single-photon emission computed tomography; CI, confidence interval; T1, T1-weighted imaging; T2, T2-weighted imaging; FLAIR, fluid-attenuated inversion recovery; AUC, area under the curve; *Italics* = biomarker metric or outcome measure dichotomized or categorized; magnitude, association: red = small, yellow = moderate, green = large; magnitude, discrimination/classification: red = poor, yellow = fair, green = excellent; significance: red = non-significant, green = significant.
† Subgroup with intact CST
‡ Subgroup with interrupted CST
§ Adjusted for age
|| Adjusted for baseline MI LL
¶ Model developed by deep neural network algorithm.

**Table 4d.**
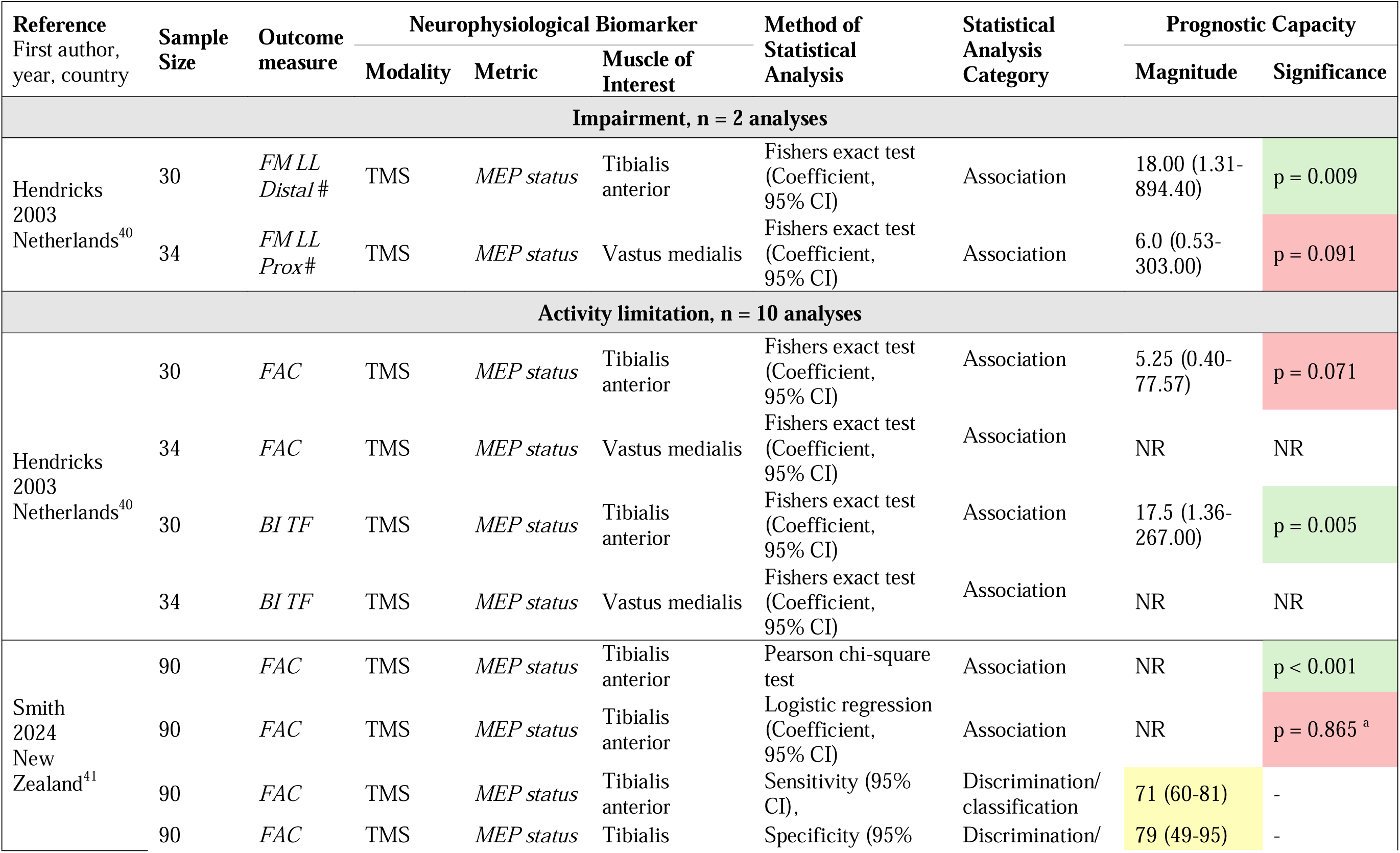
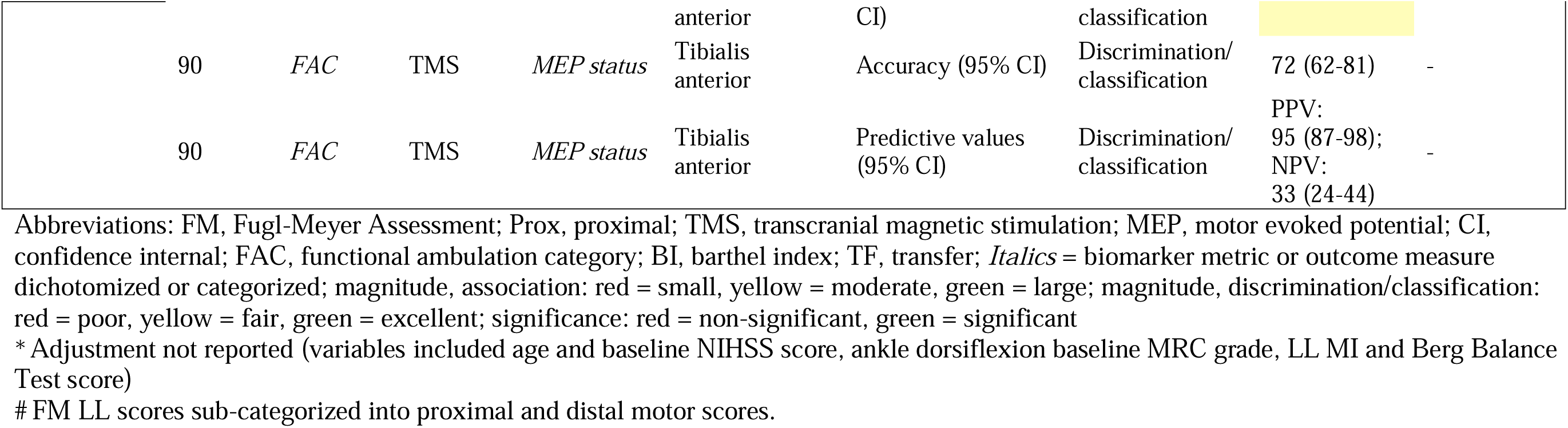
Late Subacute: Neurophysiological Biomarker Performance.

**Table 4e.** Chronic: Neuroimaging Biomarker Performance.

| Reference<br>First author,<br>year, country | Sample<br>Size | Outcome<br>measure | Neuroimaging Biomarker |  |  | Method of<br>Statistical<br>Analysis | Statistical<br>Analysis<br>Category | Prognostic Capacity |  |
| --- | --- | --- | --- | --- | --- | --- | --- | --- | --- |
|  |  |  | Modality | Metric | Tract of Interest |  |  | Magnitude | Significance |
| Activity limitation, n = 6 analyses |  |  |  |  |  |  |  |  |  |
| Jang<br>2022<br>Korea<sup>31</sup> | 30 | FAC | DTI | FA | Corticoreticulo-<br>spinal tract | Pearson<br>correlation (r) | Association | 0.295 | p = 0.11 |
|  | 30 | FAC | DTI | FA | Niagrastriatal tract | Pearson<br>correlation (r) | Association | 0.228 | p = 0.23 |
|  | 30 | FAC | DTI | TV | Corticoreticulo-<br>spinal tract | Pearson<br>correlation (r) | Association | 0.821 | p = 0.00 |
|  | 30 | FAC | DTI | TV | Nigrostriatal tract | Pearson<br>correlation (r) | Association | 0.786 | p = 0.00 |
|  | 30 | FAC | DTI | TV | Corticoreticulo-<br>spinal tract | Linear regression<br>(Coefficient,<br>95% CI) | Association | 0.532 (NR)<br>** | p = 0.00 |
|  | 30 | FAC | DTI | TV | Nigrostriatal tract | Linear regression<br>(Coefficient,<br>95% CI) | Association | 0.362 (NR)<br>†† | p = 0.04 |

Abbreviations: FAC, functional ambulation category; DTI, diffusion tensor imaging; FA, fractional anisotropy; r, Pearson Correlation Coefficient; TV, tract volume; NR, not reported; *Italics* = biomarker metric or outcome measure dichotomized or categorized; magnitude, association: red = small, yellow = moderate, green = large; significance: red = non-significant, green = significant
\*\* Adjusted for Nigrostriatal tract TV
†† Adjusted for Corticoreticulospinal tract

**Table 4f.**
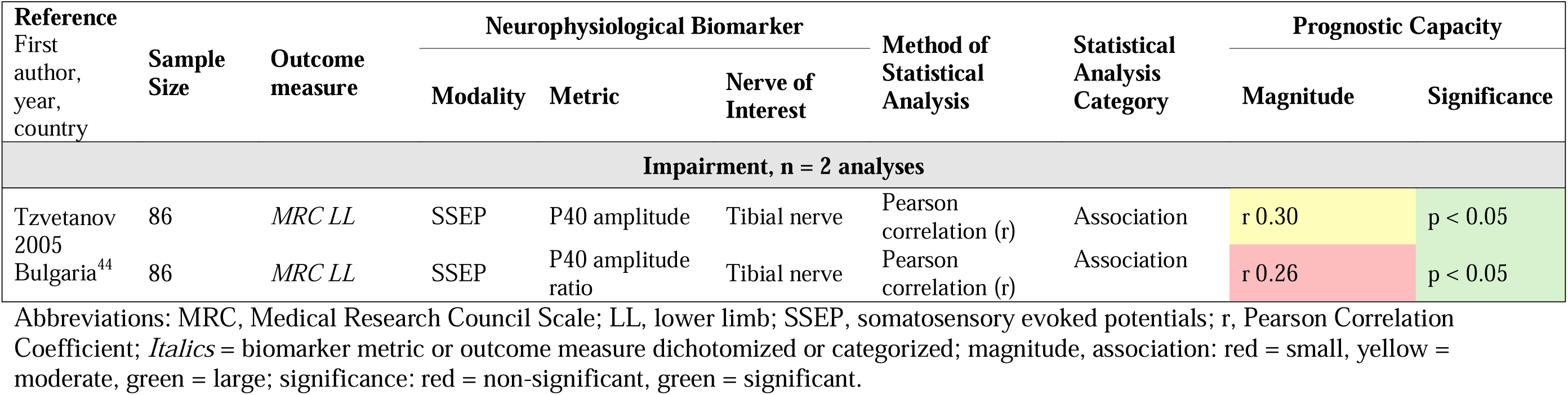
Chronic: Neurophysiological Biomarker Performance.

The following sections summarize the prognostic capacity of each biomarker by statistical analysis category. Results are grouped into neuroimaging and neurophysiological biomarkers.

### Associative Analyses

Most biomarker analyses (n=76, 88.4%) used an associative statistical approach to evaluate prognostic capacity, such as a correlation analyses (Spearman’s correlation; n=17, 19.1%) or regression analysis (logistic regression; n=16, 18.1%).

### Neuroimaging Biomarkers

#### Corticospinal Tract Structural Integrity

Six studies (n=173 participants) investigated CST structural integrity using various neuroimaging metrics, including FA, apparent diffusion coefficient (ADC), fiber number, tract area, injury grade, and lesion load. Each metric and a summary of findings are presented below.

FA parameters (n=9 analyses from five studies, including 115 participants): In the early subacute phase six analyses reported large or significant associations with lower limb motor function^25,28,29^ and independent walking ability^25^, while one found a non-significant association^30^. In the late subacute phase, two analyses reported small associations for lower limb motor function and independent walking ability^31^.

ADC parameters and lesion load (n=7 analyses from four studies, including 114 participants): All analyses reported small or non-significant associations with lower limb motor function^48,55,57^ and independent walking ability^25,30^ in the early subacute phase.

Injury grade (n=1 analysis from one study, including 28 participants): One analysis reported a large association with lower limb motor function in the early subacute phase^32^.

Fiber number and tract area (n=4 analyses from one study, including 31 participants): All analyses demonstrated moderate associations with lower limb motor function and independent walking ability in the late subacute phase^26^.

#### Non-Corticospinal Tract Structural Integrity

Two studies (including n=60 participants) examined the structural integrity of non-corticospinal tracts using FA, tract volume or lesion load. Each tract and a summary of findings are presented below.

Corticoreticulospinal and nigrostriatal tracts (n=6 analyses from one study, including 30 participants): Tract volume (n=4 analyses) demonstrated large associations with independent walking ability measured in the chronic phase, particularly for the corticoreticulospinal tract. FA (n=2 analyses) showed only small associations for both tracts^31^.

Sensorimotor tract (n=1 analysis from one study, including 30 participants): Lesion load was non-significantly associated with independent walking ability in the early-subacute phase^30^.

#### Lesion Characteristics

Eight studies (including n=311 participants) examined lesion location and size. Magnitudes were rarely reported (5.3% of lesion location analyses and 20% of lesion size analyses). This limited reporting of magnitudes constrains interpretation of their prognostic value. Each metric and a summary of findings are presented below.

Lesion location (n=19 analyses from 4 studies, including 151 participants): Only one analysis reported a measure of magnitude^33^, and two did not report any prognostic capacity results^33^. Results for outcomes measured in the early subacute phase were inconsistent: most analyses demonstrated non-significant associations with lower limb motor function (n=5)^34,35^ and independent walking ability (n=2)^34^, while a subset reported significant associations (n=1 and n=3, respectively)^33,36^. In the late subacute phase, all analyses for lower limb motor function (n=3) were non-significant^35^, whereas those assessing independent walking ability (n=3), reported significant associations^36^.

Lesion size (n=5 analyses from five studies, including 238 participants): All analyses were in relation to outcomes in the early subacute phase. Most reported non-significant associations with lower limb motor function (n=2 analyses)^28,29^ and independent walking ability (n=1 analysis)^37^, while one analysis found a large association with lower limb motor function^32^.

#### Regional Brain Characteristics

Three studies (including n=155 participants) examined measures of regional and network-level brain characteristics. Metrics included white matter hyperintensity severity, regional cerebral perfusion and interhemispheric functional connectivity. Each metric and a summary of findings are presented below.

Fazekas scale (n=3 analyses from one study, including 70 participants): Two analyses of white matter hyperintensity severity showed significant associations with independent walking ability in the late subacute phase, including one adjusted for baseline lower limb strength^38^. However, when adjusted for age, the association was no longer significant.

Hypoperfusion (n=3 analyses from one study, including 43 participants): Among participants with corona radiata infarct, reduced regional cerebral blood flow, measured using single-photon emission computed tomography, was significantly associated with independent walking ability in the late subacute phase^39^. Specifically, significant results were reported for the ipsilesional thalamus when the CST was preserved, and the contralesional cerebellum when CST was disrupted.

Interhemispheric homotopic functional connectivity (n=1 analysis from one study, including 42 participants): This study used resting state functional MRI to assess interhemispheric functional connectivity^28^ and reported no significant association with lower limb motor function in the early subacute phase.

### Neurophysiological Biomarkers

Four studies (n=235 participants) examined the associative capacity of neurophysiological biomarkers of functional corticospinal and somatosensory tract integrity. This included MEP status and somatosensory evoked potential (SSEP) parameters. Each metric and a summary of findings are presented below.

MEP status (n=12 analyses from three studies, including 149 participants): Most analyses reported non-significant associations with lower limb motor function and independent walking ability (n=6)^30,40–42^ across the early and late subacute phases. Two analyses found significant associations with independent waking ability, though this was not maintained after adjustment for age, stroke severity and baseline lower limb strength and balance^41^.

SSEP parameters (n=5 analyses from 1 study, including 86 participants): In the early subacute phase, three analyses reported inconsistent associations with lower limb motor function, ranging from small to large^43^. In the chronic phase, two analyses reported small and moderate associations^44^.

### Discrimination/Classification Analyses

Nine biomarker analyses (10.5%) used a discrimination/classification statistical approach to evaluate prognostic capacity. One neuroimaging analysis used ROC analysis to evaluate an MRI-based deep neural network algorithm; however, the AUC was not reported^45^. Seven analyses from a single study (n=90 participants) assessed the ability of MEP status to classify independent walking ability in the early and late subacute phases^48^, with four reporting fair sensitivity and specificity. Accuracy and predictive values were also reported (n=2 analyses) but not rated for prognostic capacity.

### Validation Analysis

Only one biomarker analysis applied validation methods to evaluate prognostic capacity. This analysis (n=370 participants) internally validated an MRI-based deep neural network model which demonstrated excellent discrimination of independent walking ability at the late subacute phase^45^.

## Discussion

This systematic review synthesized current evidence on the prognostic capacity of early neuroimaging and neurophysiological biomarkers for mobility outcomes post-stroke. Most analyses of early biomarkers included in this review remained at the association stage, and results were generally small or non-significant. Collectively, the evidence base is insufficient to identify early biomarkers with potential for clinical translation. These findings address gaps of previous systematic reviews^6–8^ to advance our understanding of early biomarkers of mobility recovery.

Corticospinal tract integrity has long been considered a key indicator of mobility recovery post-stroke^5^. The findings from this review challenge this assumption. Despite frequent investigation, early measures of structural and functional CST integrity showed limited prognostic capacity for mobility outcomes across the early to late subacute phases. Structural biomarkers of CST, including FA, ADC and lesion load, showed inconsistent and mostly small or non-significant results. While CST FA showed some promising associations, this must be interpreted with caution given the small sample sizes and heterogeneity^25,28,29^. Similarly, functional CST integrity, assessed via MEP status, was only weakly associated with lower limb function or independent walking. In the few analyses with significant associations, these did not hold after adjusting for demographic and clinical factors. In the only study that assessed the classification performance of MEP status, sensitivity and specificity were fair alongside poor negative predictive values, raising concerns about the risk of misclassifying patients as unlikely to recover^41^. Such misclassifications could result in overly pessimistic prognoses and inappropriate restriction of access to rehabilitation. While these findings are based on small studies with moderate to high risk of bias, the overall evidence suggests that early CST integrity biomarkers currently offer limited prognostic value and should not be used in isolation to guide post-stroke mobility prognostication.

Lesion characteristics, including lesion location and size, have been frequently examined as candidate neuroimaging biomarkers. Their prognostic value remains unclear due to inconsistent associative findings and limited reporting of magnitudes. Further research using standardised discrimination or classification based statistical approaches, and consistent reporting is needed to clarify their prognostic utility.

More recent research is exploring emerging biomarkers that may better reflect the complexity of mobility recovery post-stroke. These include indicators of the broader interplay of multiple brain regions^39^, bilateral descending pathways to the trunk and proximal lower limb muscles, or spinal locomotor circuits^31^. While preliminary associative findings are promising, these studies are limited by small sample sizes and selective populations^31,39^. Further investigation in larger, more representative cohorts is needed to confirm their prognostic utility. As research in this area expands, it may offer more comprehensive insights into the neural substrates that support mobility recovery and inform the development of more robust prognostic models.

Only one study internally validated a prognostic model, which marks an initial step toward clinical translation. Shin et al.^45^ developed a deep neural network algorithm using MRI data to distinguish between individuals likely or unlikely to achieve independent walking at six months post-stroke. As the largest study in this review, the model demonstrated excellent discriminative performance via internal validation using a random split-sample. Areas of moderate to high risk of bias should be considered before progressing to an external validation study to guide clinical translation. Practical barriers, including technological infrastructure and workforce expertise, may limit immediate clinical implementation. Nonetheless, as machine learning approaches become more accessible, this may represent a promising direction for future prognostic research.

There were substantial methodological limitations of the studies included in this review, which constrains the conclusions that can be drawn. A key limitation was the inadequate reporting of essential prognostic indicators, such as effect magnitudes, which restricts interpretation of clinical relevance. Small sample sizes and moderate to high risk of bias were common, further reducing confidence in reported associations. Underreporting of key clinical factors, such as baseline stroke severity and acute medical interventions, also limits assessment of whether differences in study populations contributed to variability in findings. Finally, inconsistent use of statistical methods, outcome measures and follow-up timepoints, reduces comparability and limits synthesis across studies.

Given these methodological limitations, and the practical barriers to biomarker collection (e.g., cost, workforce demands and accessibility to equipment) particularly in non-metropolitan settings, it may be more feasible to shift focus toward the refinement and implementation of validated clinical tools. Such tools may offer a more immediate path to improving early prognostication, while biomarker research continues to evolve. Early clinical measures such as trunk control^2,46^, balance^47^ and lower limb strength^2,46,47^, combined with demographic and stroke-related factors like age^1,2^ and stroke severity^1^ have been found to be associated with mobility outcomes. Prognostic tools such as the Time to Walking Independently after Stroke (TWIST)^47^ and Early Prediction of Functional Outcome after Stroke (EPOS)^46^ have demonstrated acceptable prognostic performance. Further research involving larger, non-selective samples and international external validation is needed to confirm generalizability, and ongoing studies are expected to address this gap^48^.

This review was rigorous, including a comprehensive search strategy, two independent reviewers at all stages, and inclusion of non-English studies. There were some limitations. Meta-analysis was not possible due to substantial heterogeneity in biomarkers, outcome measures, follow-up time points, statistical methods and reporting across studies. Despite repeated contact, few authors provided missing or incomplete results (e.g., magnitudes). Lastly, many studies had areas of moderate to high risk of bias, which constrain the strength of conclusions.

## Conclusion

This review specifically focused on biomarkers measured within 14 days post-stroke, reflecting the clinical priority of early prognostication to guide timely decision-making. Current evidence indicates that early neuroimaging and neurophysiological biomarkers offer limited prognostic utility for post-stroke mobility. Most studies were underpowered and lacked methodological rigor. Despite the 2017 SRRR recommendations highlighting the need for high-quality research in this area, progress has been limited as only six studies included were published since 2017^28,31,38,39,41,45^. To advance the field, coordinated international collaboration with harmonized methodologies, standardised statistical reporting and consistent use of SRRR recommended timepoints^18^ and outcome measures^49^ is needed. The development of integrative prognostic tools that combine clinical and biomarker measures may prove valuable in sub-populations with greater prognostic uncertainty. Our review emphasizes a growing recognition of the limitations of isolated biomarkers and the need for comprehensive, systems-level approaches to prognostication.

## Nonstandard Abbreviations and Acronyms

**In text and in tables:**

(ADC): Apparent diffusion coefficient
(AUC): Area under the curve
(BI): Barthel index
(CT): Computed tomography
(CI): Confidence interval
(CST): Corticospinal tract
(DTI): Diffusion tensor imaging
(FAAI): FA asymmetry index
(FN): Fiber number
(FLAIR): Fluid-attenuated inversion recovery
(FA): Fractional anisotropy
(FM): Fugl Meyer Assessment
(FAC): Functional ambulation category
(FIM): Functional independence measure
Interhemispheric functional connectivity: 
(ICARS): International Cooperative Ataxia Rating Scale
(LL): Lower limb
(MRI): Magnetic resonance imaging
(MRC): Medical Research Council Scale
(mRS): Modified Rankin scale
(MEP): Motor evoked potential status
(MI): Motricity index
(NIHSS): National Institutes of Health Stroke Scale
(QUIPS): Quality in Prognostic Factor Studies
(ROC): Receiver Operating Characteristic
(rsfMRI): Resting state functional MRI
(SSS): Scandinavian Stroke Scale
(SPECT): Single-photon emission computed tomography
(SSEP): Somatosensory evoked potential
(SRRR): Stroke Recovery and Rehabilitation Roundtable
(T1): T1-weighted imaging
(T2): T2-weighted imaging
(TV): Tract volume
(TMS): Transcranial magnetic stimulation
(TF): Transfer

## Acknowledgements

None.

## Supplemental Material

MEDLINE Search Strategy.

