## Supplemental Table S1 for "A systematic review of early neuroimaging and neurophysiological biomarkers for post-stroke mobility prognostication"

**Table S1. MEDLINE Search Strategy**

| MEDLINE | | | |
| --- | --- | --- | --- |
| Population | **Intervention** | **Outcome** | |
| Stroke survivors | **Neuroimaging and**  **neurophysiological biomarkers** | **Upper limb function** | **Mobility** |
| 1. exp cerebrovascular disorders/ 2. exp stroke/ 3. (stroke$ or cva$ or cerebrovascular or cerebral vascular).mp. 4. (poststroke or post-stroke or cerebrovasc$ or brain vasc$ or cerebral vasc$ or apoplex$ or SAH).mp. 5. ((brain$ or cerebr$ or cerebell$ or intracran$ or intracerebral or vertebrobasilar) adj5 (isch?emi$ or infarct$ or thrombo$ or emboli$ or occlus$)).mp. 6. ((brain$ or cerebr$ or cerebell$ or intracerebral or intracranial or subarachnoid) adj5 (haemorrhage$ or hemorrhage$ or haematoma$ or hematoma$ or bleed$)).mp. 7. Hemiplegia/ or Hemiparesis/ or Paresis/ 8. (hemipleg$ or hemipar$ or paresis or paretic).mp. | 9. Functional connect$.mp. 10. Neurophys$.mp. 11. exp Transcranial Magnetic Stimulation/ 12. (TMS or Transcranial magnetic stimulation or MEP or Motor evoked potential$).mp. 13. exp Neuroimaging/ 14. (Neuroimag$ or Neuro Imag$ or Brain Imag$ or Brain map$).mp. 15. (fMRI or functional MRI or functional magnetic resonance imag$).mp. 16. ((Resting state) adj3 (network$ or brain map$ or magnetic resonance imag$ or MRI) or rsfMRI or rsMRI).mp. 17. exp electroencephalography/ 18. (EEG or Electroencephalography).mp. 19. Structural connect$.mp. 20. exp Tomography, X-Ray Computed/ 21. (CT or Computed tomography).mp. 22. exp Magnetic Resonance Imaging/ 23. (MRI or NMRI or Magnetic Resonance imag$).mp. 24. (T1-weighted or T1 weighted or T1WI or T1 or T2-weighted or T2weighted or T2WI or T2 or T2*-weighted or T2*WI or T2* or T2*-Gradient Echo or T2*-GRE or Fluid attenuated inversion recovery or FLAIR).mp. 25. (Diffusion Tensor$ or DTI or DTI-MRI or Tractography).mp. 26. (Diffusion Weighted Imag$ or Diffusion imag$ or DWI).mp. | 27. exp Upper Extremity/ 28. (upper limb$ or upper extremit$ or arm or hand or shoulder or elbow or forearm or wrist or finger).mp. | 29. exp gait/ or walking/ or locomotion/ 30. exp mobility limitation/ 31. (walk$ or gait$ or mobil$ or ambulat$ or locomot$ or balanc$).mp. 32. exp lower extremity/ 33. (lower limb$ or lower extremit$ or leg or pelvi$ or hip or knee or ankle or foot or feet or toe).mp. |
| 34. 1 or 2 or 3 or 4 or 5 or 6 or 7 or 8 | 35. 9 or 10 or 11 or 12 or 13 or 14 or 15 or 16 or 17 or 18 or 19 or 20 or 21 or 22 or 23 or 24 or 25 or 26 | 36. 27 or 28 or 29 or 30 or 31 or 32 or 33 | |
| 37. (Validat$ or Predict$ or Rule$ or (Predict$ and (Outcome$ or Risk$ or Model$)) or ((History or Variable$ or Criteria or Scor$ or Characteristic$ or Finding$ or Factor$) and (Predict$ or Model$ or Decision$ or Identif$ or Prognos$))).mp. or (Decision$.mp. and ((Model$ or Clinical$).mp. or logistic Models/)) or ((Prognostic and (History or Variable$ or Criteria or Scor$ or Characteristic$ or Finding$ or Factor$ or Model$)) or (Associat$ or Correlat$)).mp. | | | |
| 38. 34 and 35 and 36 and 37  39. Limit 38 to Humans | | | |
